# Modeling Biases in SARS-CoV-2 infections Prediction using Genome Copies Concentration in Wastewater

**DOI:** 10.1101/2023.03.06.23286832

**Authors:** Mattia Mattei, Rosa M. Pinto, Susana Guix, Albert Bosch, Alex Arenas

## Abstract

**Background:** SARS-CoV-2, the virus responsible for the COVID-19 pandemic, can be detected in stool samples and subsequently shed in the sewage system. The field of Wastewater-based epidemiology (WBE) aims to use this valuable source of data for epidemiological surveillance, as it has the potential to identify unreported infections and to anticipate the need for diagnostic tests.

**Objectives:** The objectives of this study were to analyze the absolute concentration of genome copies of SARS-CoV-2 shed in Catalonia’s wastewater during the Omicron peak in January 2022, and to develop a mathematical model capable of using wastewater data to estimate the actual number of infections and the temporal relationship between reported and unreported infections.

**Methods:** We collected twenty-four-hour composite 1-liter samples of wastewater from 16 wastewater treatment plants (WWTPs) in Catalonia on a weekly basis. We incorporated this data into a compartmental epidemiological model that distinguishes between reported and unreported infections and uses a convolution process to estimate the genome copies shed in sewage.

**Results:** The 16 WWTPs showed an average correlation of 0.88 *±* 0.08 (ranging from 0.96 to 0.71) and an average delay of 8.7 *±* 5.4 days (ranging from 0 to 20 days). Our model estimates that about 53% of the population in our study had been infected during the period under investigation, compared to the 19% of cases that were detected. This under-reporting was especially high between November and December 2021, with values up to 10. Our model also allowed us to estimate the maximum quantity of genome copies shed in a gram of feces by an infected individual, which ranged from 4.15 *×* 10^7^ *gc/g* to 1.33 *×* 10^8^ *gc/g*.

**Discussion:** Although wastewater data can be affected by uncertainties and may be subject to fluctuations, it can provide useful insights into the current trend of an epidemic. As a complementary tool, WBE can help account for unreported infections and anticipate the need for diagnostic tests, particularly when testing rates are affected by human behavior-related biases.

## 1 Introduction

The emergence of the SARS-CoV-2 coronavirus in 2019 has resulted in a global pandemic, which has led to over 600 million infections and 6 million deaths worldwide. Epidemiological data has played a crucial role in monitoring the spread of the epidemic, with clinical testing via reverse transcription quantitative polymerase chain reaction (Rt-qPCR) on nasopharyngeal swabs being the primary method. However, limitations and biases exist in any epidemiological indicator, particularly in the case of PCR testing, which relies on voluntary participation and may only capture individuals more likely to be infected. Given the high number of asymptomatic and subclinical infections, this biased testing process can significantly impact estimations. Although hospitalizations and deaths are less susceptible to bias, they may not be ideal for real-time forecasting due to their lagged estimations.

Wastewater-based epidemiology (WBE), i.e. the surveillance of epidemic spreading through the analysis of virus concentration in wastewater plants, is therefore presenting itself as a potential complementary tool to clinical testing, and it is gaining more and more attention among the mathematical modellers. The concept of WBE centers around the knowledge that SARS-CoV-2 RNA can be detected in stool samples excreted by human bodies [1–3], and then shed in the sewage system. Therefore, daily sampling of SARS-CoV 2 RNA in wastewater would provide information similar to that from daily random testing of thousands of individuals in a community [4], but not distinguishing between symptomatic, asymptomatic or presymptomatic people as long as they develop viral RNA in their feces. The interest of the WBE relays on two main aspects: wastewater data can potentially account the unreported cases, and they can also represent an estimate in advanced over time respect to diagnostic tests. Consequently, WBE is envisaged to become the most important non-invasive diagnostic tool of the epidemics in a population.

The concept of wastewater epidemiology has frequently been referred to as a “leading indicator” of reported cases [5], although there is often a time delay between the two measures [6–9]. The extent of the lead time provided by sewage data varies significantly in the literature, ranging from a few days to up to two weeks [10]. The duration of the time delay between wastewater estimates and reported cases is influenced by various factors, including the characteristics of the health system such as the availability and distribution of diagnostic tests, and the time required to obtain test results [5]. However, it is important to note that wastewater estimates are subject to considerable uncertainty, which is attributable to several factors. Firstly, our understanding of the shedding process is limited. As detailed in section 2.2, the quantity of SARS-CoV-2 shed in feces and its temporal profile exhibit considerable inter-individual variability, and clinical studies have reported a wide range of results. Moreover, it remains unclear whether the onset of viral shedding in feces precedes or coincides with the onset of symptoms, given that most existing clinical studies have been conducted in hospitalized patients.

Secondly, the virus within the sewage system is subject to various “random” factors, such as dilution with the daily water flow, temperature, possible interactions with chemical agents or other substances, and environmental factors like rain. The features of the sewage systems, such as the travel time from households to treatment plants, can also have a significant impact on measurements. Lastly, the experimental process for extracting data on genetic copy concentration from sewage is not without challenges, and measurement errors should always be taken into consideration.

Upon considering the previous explanations, a fundamental question arises as to how to mathematically quantify the biases between the genome copies concentration in wastewater, the reported cases, and the actual incidence in any given area. To this end, we conducted an analysis of data pertaining to the absolute concentrations of the SARS-CoV-2 gene biomarker N1 in weekly wastewater samples collected from 16 wastewater treatment plants (WWTP) in Catalonia, Spain, during the period spanning October 2021 to March 2022. The data was sourced from the Catalan Surveillance Network of SARS-CoV-2 in Sewage (https://sarsaigua.icra.cat/). Initially, we examined the time delay and statistical correlation between wastewater data and reported cases at each WWTP. The data pertaining to reported cases was obtained from the official website of the Catalonia government (Generalitat de Catalunya https://analisi.transparenciacatalunya.cat/browse?q=covid&sortBy=relevance). Subsequently, we proposed a model that incorporates a time-varying rate of unreported cases to explain the observed delays and, in general, the heterogeneity of outcomes reported in the literature on the subject.

The principal outcomes of our study are twofold: Firstly, our analysis of wastewater data in the Catalonia region reveals a markedly high correlation with reported cases, with a mean Pearson correlation of 0.9, and an average 9-day advance in anticipating trends in reported cases, but with variability ranging from 0 to 20 days. Secondly, the proposed model enables us to successfully link wastewater data with temporal dynamics of the reported cases during the same period, and provides estimates of the actual prevalence of infection and parameters of interest in the context of wastewater-based epidemiology.

## 2 Methods

### 2.1 Wastewater sampling

The study involved the weekly collection of 1-liter composite samples of influent wastewater from 16 wastewater treatment plants (WWTPs) (Table 1) in the region of Catalonia, Spain. The samples were collected over a period of six months, from October 2021 to March 2022. The WWTPs selected covered a population of approximately 2,514,618 inhabitants, which represents around 31% of the total Catalan population. The samples were transported in a portable icebox at a temperature range of 0°-4°C and were analyzed the day after concentration. The wastewater samples were collected within the framework of the Catalan Surveillance Network of SARS-CoV-2 in Sewage (https://sarsaigua.icra.cat/; http://doi.org/10.5281/zenodo.4147073).

**Table 1:**
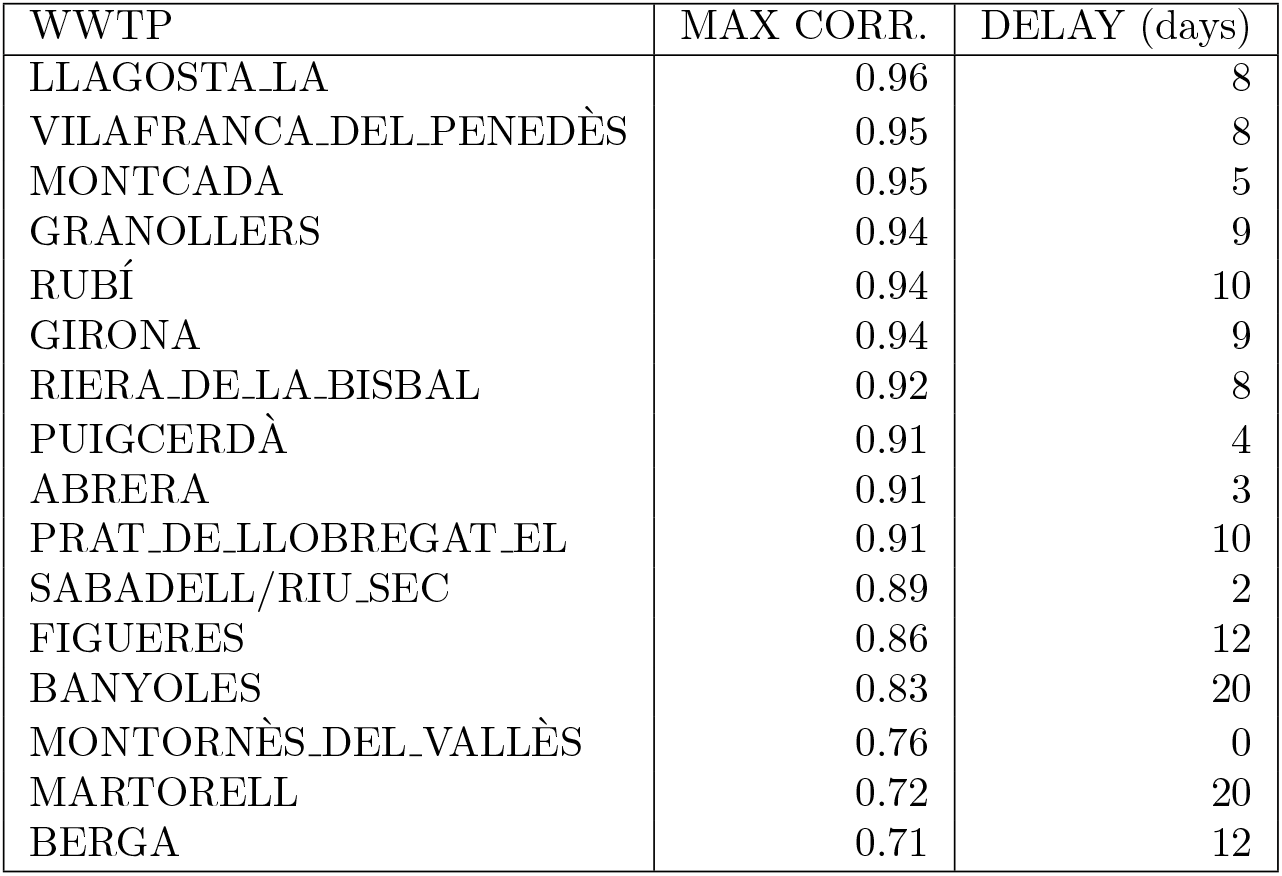
The maximum Pearson correlations and the corresponding delays between sewage data and reported cases for each WWTP. For each of the 16 wastewater plants listed in the left column we measured the Pearson correlation between genome copies concentrations data linearly interpolated and 7-days averaged daily reported cases in the corresponding served municipalities. We performed the analysis shifting back in time reported cases from 0 to 20 days. We interpreted as *delay* the shift at which we reached maximum correlation.

### 2.2 Wastewater concentration

Wastewater samples were concentrated by using the aluminum hydroxide adsorption-precipitation method, as described in previous studies (Randazzo et al., 2020 [11]; Wallis-Melnick, 1967 [12]). Two hundred milliliters of wastewater were concentrated to a final volume of 1-5 milliliters of phosphate-buffered saline (PBS). To ensure the accuracy of the concentration process, 1.5 × 10^6^ TCID50 units of the attenuated PUR46-MAD strain of the Transmissible Gastroenteritis Enteric Virus (TGEV) (Moreno et al., 2008 [13]) were seeded into each sample prior to the concentration step.

### 2.3 Nucleic acid extraction

To extract nucleic acids, 300 *μ*l of the concentrated samples were used and the Maxwell® RSC PureFood GMO and Authentication Kit (Promega) was employed following the manufacturer’s instructions. In each extraction run, a PBS negative control was included. To determine virus recovery, a previously described RTqPCR assay for quantification of the Transmissible Gastroenteritis Enteric Virus (TGEV) was used (Vemulapalli and GulaniSantrich, 2009 [14]). Samples with virus recovery ≥ 1% were deemed acceptable. Recoveries varied from 1% to 98% with an average of 24% ± 18%.

### 2.4 RTqPCR assays

Quantification of SARS-CoV-2 RNA in sewage samples was based on the N1 assay (US-CDC 2020), which targets a fragment of the nucleocapsid gene. We employed the PrimeScript™ One Step RT-PCR Kit (Takara Bio, USA) and a CFX96 BioRad instrument.

### 2.5 Convolution description of viral shedding

Convolution operations represent the most appropriate approach to mathematically model the relationships between genome copy concentration, reported cases, and actual infection prevalence. Convolution is a mathematical method that involves the combination of two functions to produce a third function that describes how one function modifies the other. The resulting function is defined as the convolution of the two input functions. In essence, the procedure involves sliding one function over the other, multiplying the overlapping portions of the two functions, and integrating the product over the entire variable range to generate a novel function that portrays the interplay between the two functions.

The virus concentration in sewage can be modeled as a function of the number of infected individuals in the serviced area and the time since they became infected, given a specific profile of the quantity of virus shed in feces over time.

There is a general consensus in the scientific literature on certain characteristics of the virus shedding profile: long duration, exponential decay, and peak around symptom onset. Wu et al. [15] reported that SARS-CoV-2 RNA can be detected in feces for a mean of 11.2 days after respiratory tract samples test negative (up to 5 weeks); Zhang et al. [16] found a median fecal shedding duration of 22 days. Wolfel et al. [1] observed RNA-positive stool samples for over 3 weeks without symptoms, with peak viral RNA likely occurring during the first week of symptoms. The timing of shedding onset relative to symptom onset remains debated due to lack of clinical data on exposed individuals, but Hoffman et al. [17] constrained the latter part of the shedding profile with a fast exponential decay. Miura et al. [18] successfully tested the model proposed by Teunis et al. [19] for norovirus shedding, to SARS-CoV-2 clinical data, which accounts for both exponential rise and decay.

Wrapping up, a description that accurately captures the essential characteristics of the viral shedding profile is a gamma distribution, as reported in previous studies such as Huisman et al. [20] and Fernandez-Cassi et al. [21]. Specifically, Huisman et al. [20] used data on the incubation period and gastrointestinal shedding following symptoms onset to model the shedding profile as a gamma distribution with a mean of 6.7 days and a standard deviation of 7 days. We adopted this approach in our analysis.

Therefore, we modeled the quantity of genome copies at day *t* as

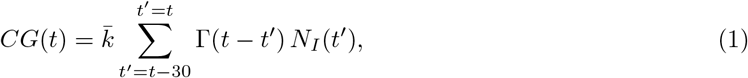

where the number of new infections at time *t*^*′*^, *N*_*I*_ (*t*^*′*^), is convoluted with the gamma distribution described above, truncated at 30 days, which tell us the quantity of genome copies per gram of feces shed at *t* − *t*^*′*^ days after the infection. The factor 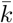 is a scale parameter and it should take in account several aspects: the degradation, *D* (defined between 0 and 1), that the shed virus may undergoes in his way to the plant (this is affected by multiple factors like water temperature, dilution, chemical reactions as well as by the time the virus spends in it), the average grams of feces produced per person *g*, the fraction of infected people shedding virus in feces *p* and the total quantity of virus shed in a gram of feces by an individual during the entire course of the infection *Q*. Therefore, similarly as in Ahmed et al. [22], we can write

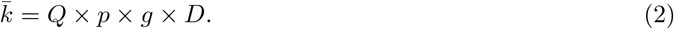

Following Chavarria-Miró et al. [23] the quantity *g* can be taken equal to 380 grams per day, based on an excretion of 30 *g* per 5.5 *kg* of body weight, assuming an average weight of Spanish population of 70 *kg* (https://www.mscbs.gob.es/estadEstudios/sanidadDatos/). The value for the fraction of infected people shedding virus in feces *p*, is quite variable in the literature, ranging from the 29% of Wang et al. [2] to the 83.3 % of the patients for Zhang et al. [16]. The review on the topic by Cheung et al. [24] suggests to consider a value equal to the 48.1%.

The value of Q is also uncertain. Several studies [1] [24] [25] [26] agree that the maximum possible shed quantity of genome copies per milliliter of stool should be around *Q*_max_ = 10^7^ gc/ml; Zhang et al. [16] indicates one order of magnitude less (*Q*_max_ = 10^5.8^ gc/ml) while Arts et al. [27] proposed a value around *Q*_max_ = 10^9^ gc/g. The dissipation process is also quite difficult to describe as affected by random factors and, in principle, it could change in time. McMahan et al. [25] proposed to use an exponential decay model which considers the effects of the water temperature and of the holding time on the virus. As stated by Weidhaas et al. [28], reported decay rates for SARS-CoV and surrogate coronaviruses in unpasteurized wastewater at 23 °C range from 0.02 to 0.143 per hour.

### 2.6 Compartmental model with a time-varying rate of reported infections

Compartmental models, specifically ordinary differential equation (ODE) models, have been the cornerstone of infectious disease modeling for over a century. These models divide the population into different compartments based on their infectious status, such as susceptible, infected, and recovered in the classical SIR model [29]. The movements of individuals between compartments are described by transition rates, which are based on the underlying biology of the disease, as well as demographic and behavioral factors. By simulating these transitions, compartmental models can be used to predict the future course of an outbreak and to evaluate the impact of different intervention strategies (Arenas et al. [30]).

In this study, we propose a variation of the Susceptible-Infected-Recovered (SIR) model, where infected individuals are divided into those who are infected but not detected (*I*_*N*_) and those who are detected and isolated (*I*_*D*_). The model is described by a system of differential equations, where the transmission rate of the disease is represented by *β*, the recovery rate by *γ*, and the total population by *N* :

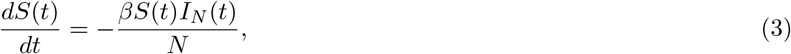

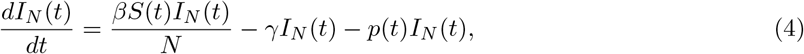

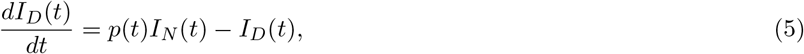

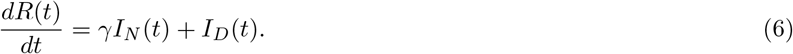

Note that we have included in the model a time-dependent probability of infected individuals being detected, represented by *p*(*t*), which is proportional to the ratio of the detected infections at time *t*:

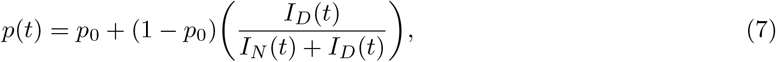

where, in case of zero detection, *p*(*t*) = *p*_0_. This equation consists of a constant part and a time-dependent one: at each time step there is a constant percentage *p*_0_ of infected that decide to get tested *unconditionally* while the rest is more sensible to the available information about the actual state of the epidemics.

This probability is influenced by factors such as changes in testing availability, policy, and implementation of Non-Pharmaceutical-Interventions (NPIs), as well as the general perception of the population about the ongoing epidemic. We assume that infected individuals, once detected, are automatically removed from the infected but not detected compartment. Moreover, we argue that *p*(*t*) is also fundamentally related to the general perception that the population has about the on-going epidemic, especially when the testing process is subministered on voluntary basis: people can be more or less willing to be tested according to their risk awareness or according to costs/benefits considerations, which clearly depend on the state of the epidemic, or better, on its *perceived* state; all the information that people have about the epidemic are enclosed in the daily reported cases. Given this and taking inspiration by several works which tried to model risk perception ([31], [32]).

Our idea is to validate this model using both wastewater data and reported cases information. The former can be generated at each time step according to equation 1, with daily new infections estimated by the system of equations above.

We argue that our model has the potential to provide insights into parameters of interest, such as 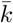 and *p*(*t*), which can justify the spectrum of delays observed between wastewater data and reported cases. This theoretical framework can provide valuable information about the dynamics of infectious diseases and can inform public health policy and decision-making.

## 3 Results

### 3.1 Statistical description

We calculated the Pearson correlation between the number of genome copies in each wastewater treatment plant (WWTP) and the 7-day averaged number of reported COVID-19 cases for each specific plant. The reported cases were shifted back from 0 to 20 days to quantify the delay between sewage data and reported cases. We analyzed the period between October 2021 and March 2022, during which the Omicron variant was spreading rapidly in Catalonia and other parts of the world. The results, summarized in Table 1 and Figure 1, showed an average correlation of 0.88±0.08 (0.96-0.71) and an average delay of 8.7±5.4 (0-20) days across the 16 WWTPs.

**Figure 1:**
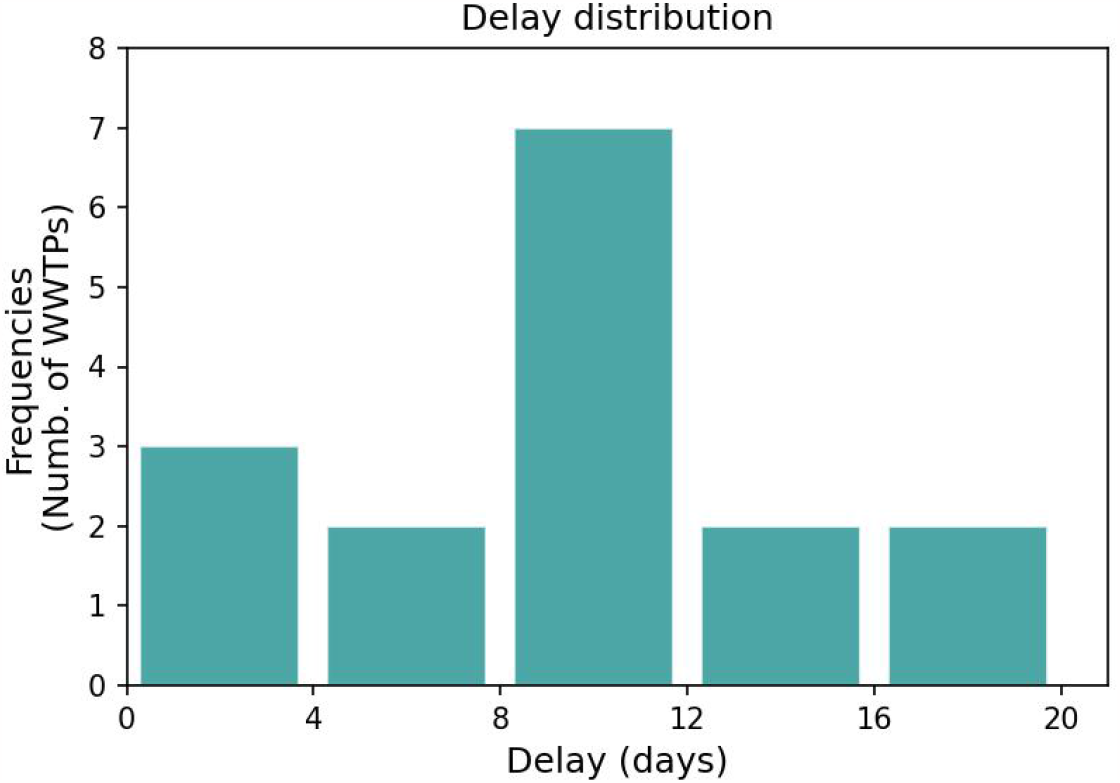
Distribution of the delays between wastewater data and reported cases curve across the 16 WWTPs analyzed.

These findings suggest that wastewater data can broadly capture the current trend of the epidemic, or at least to the extent that reported cases do. Furthermore, they seem to anticipate voluntary testing by a relevant quantity of days, more than reported in other studies. The observed delay was highly heterogeneous, ranging from 0 to 20 days, with extreme values occurring in the case of lower correlations.

Figure 2 compares the genome copies per liter averaged on all the 16 WWTPs versus the cases reported for the entire Catalonia, both normalized according to a population of 100,000 inhabitants. In this global perspective, the time-shift between the two curves is 10 days, with a correlation of 0.95. The model will provide plausible arguments to realistically explain a delay higher than expected, considering the available information about incubation period, fecal shedding, infection duration, and in general, to justify the wide range of observations in the literature.

**Figure 2:**
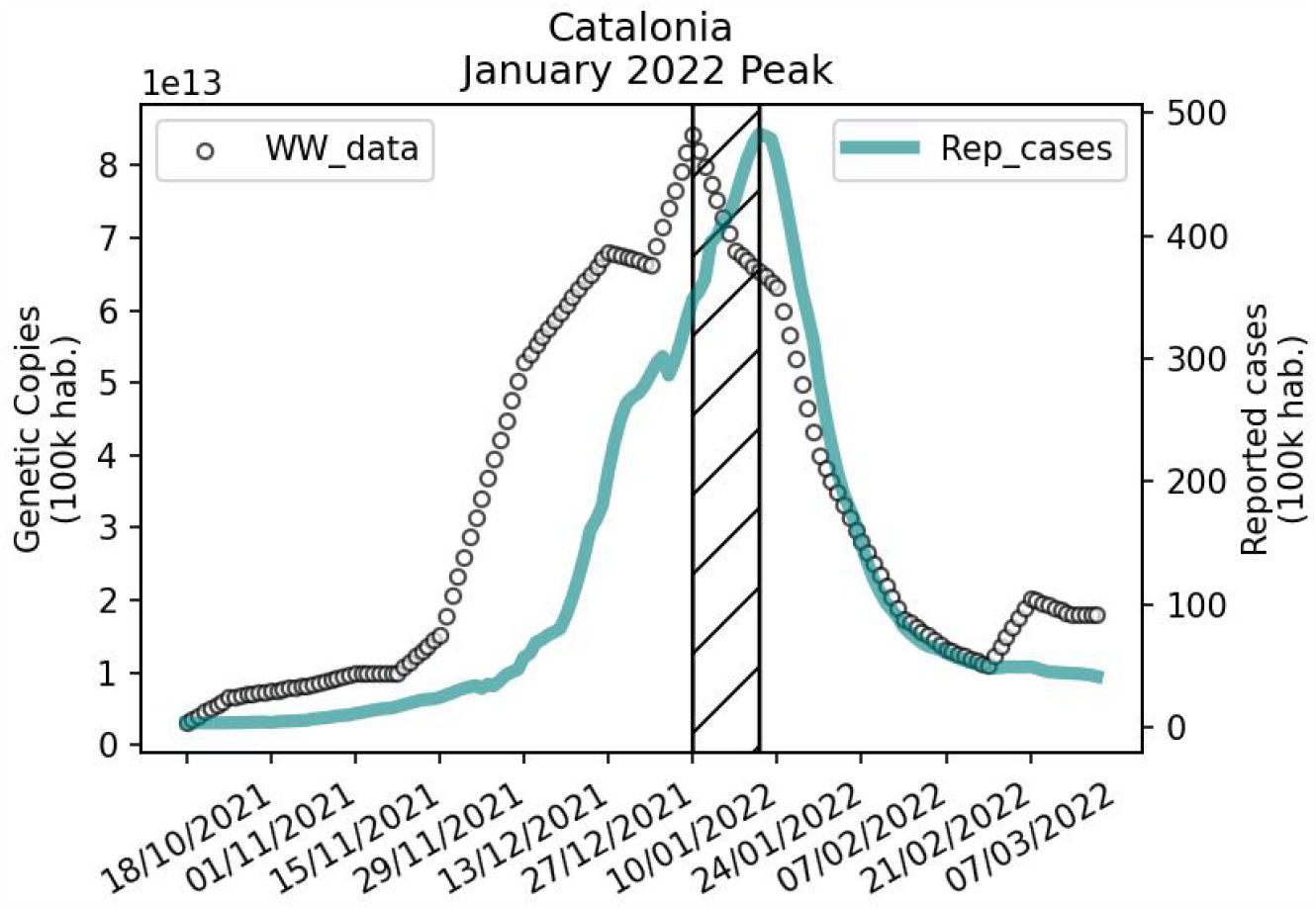
Absolute genome copies concentration averaged for all the WWTPs versus reported cases for the entire Catalonia.

### 3.2 Model calibration and validation

The model has been calibrated with real data of reported cases and genome copies concentration, averaged across all the 16 plants and normalized to 100.000 inhabitants, using Approximate Bayesian Computation (ABC) [33]. For a total time period of 152 days, we trained the model with the first 100 days and then we validated it for the remaining ones. The procedure converged yielding the posterior distributions for parameters *β, p*_0_ and 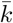 displayed in figure 3. The parameter *γ* has been chosen equal to 10 days^−1^. All the details about the parameters can be found in table 2.

**Table 2:** Parameters of the model. The parameters were estimated using Approximate Bayesian Computation (ABC). Only the recovery rate *γ* was assumed to be equal to 10 days^−1^.

| Symbol | Description | Estimates | Assignment |
| --- | --- | --- | --- |
| $\beta$ | Infectivity | 0.225 (97% CI: 0.223 - 0.227) | Calibrated |
| $\gamma$ | Recovery rate | 10 days<sup>-1</sup> | Assumed |
| $p_0$ | Initial testing rate | 0.004 (97% CI: 0.001 - 0.006) | Calibrated |
| $k$ | Scale factor for shedding process | $9.49 \times 10^{10}$ (97% CI: $9.29 \times 10^{10}$ - $9.68 \times 10^{10}$) | Calibrated |

**Figure 3:**
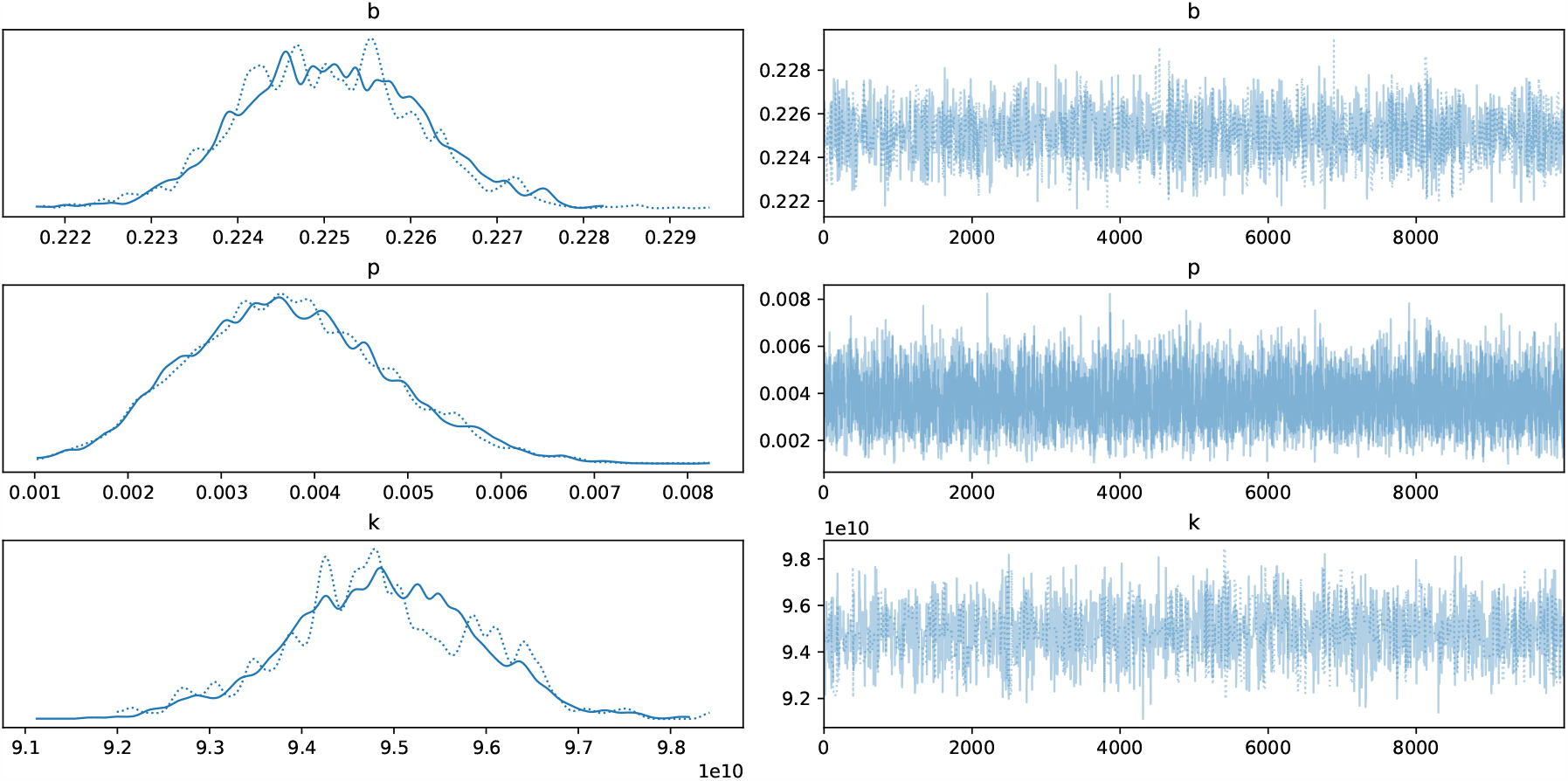
Posterior distributions provided by Approximate Bayesian Computation for parameters *β, p*_0_ and 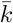 (b, p and k respectively in the figure).

Afterwards, we ran the model using the average parameter values obtained from the calibration process. The resulting epidemiological scenario is presented in Figure 4, which shows the proportions of susceptible individuals (*S*), undetected infected individuals (*I*_*N*_), detected infected individuals (*I*_*D*_), and recovered individuals (*R*).

**Figure 4:**
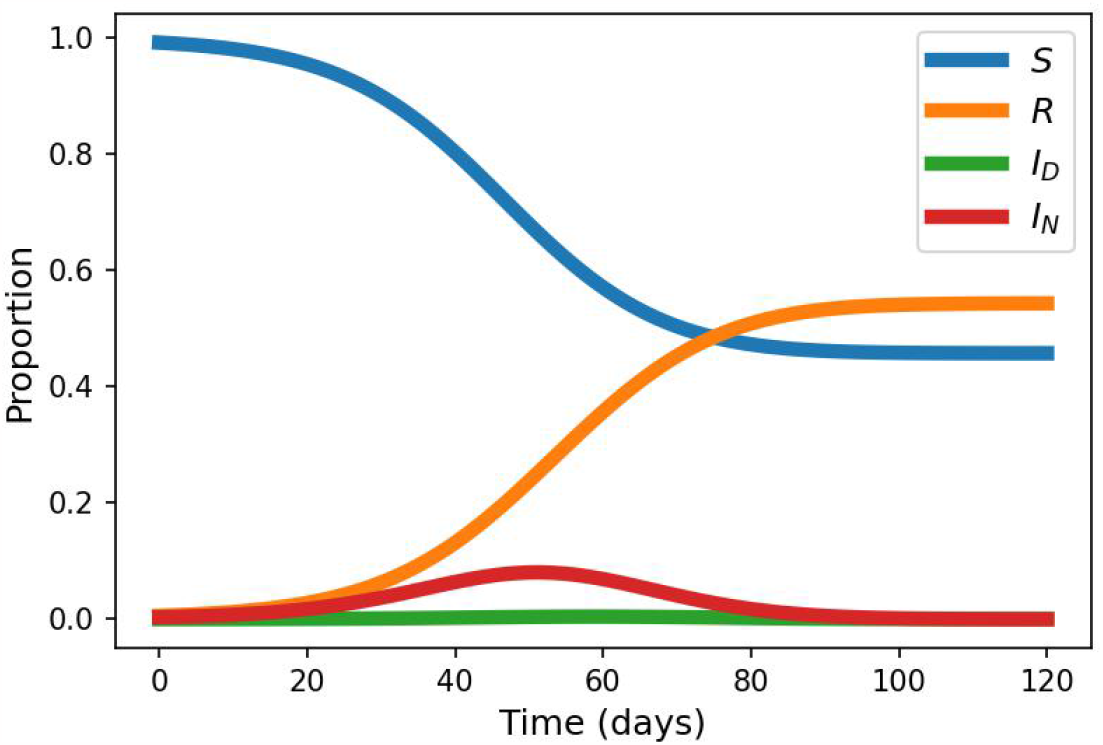
Proportions of the population being susceptible (*S*), infected not detected (*I*_*N*_), infected detected (*I*_*D*_) and recovered (*R*).

According to the model, approximately 53% of the population under study was infected during the period analyzed. Figure 5 presents a comparison between the confirmed cases data, wastewater data, and the model’s predictions in the left and right panels, respectively. The *R*^2^ statistics for reported cases and genome copies are 0.94 and 0.64, respectively.

**Figure 5:**
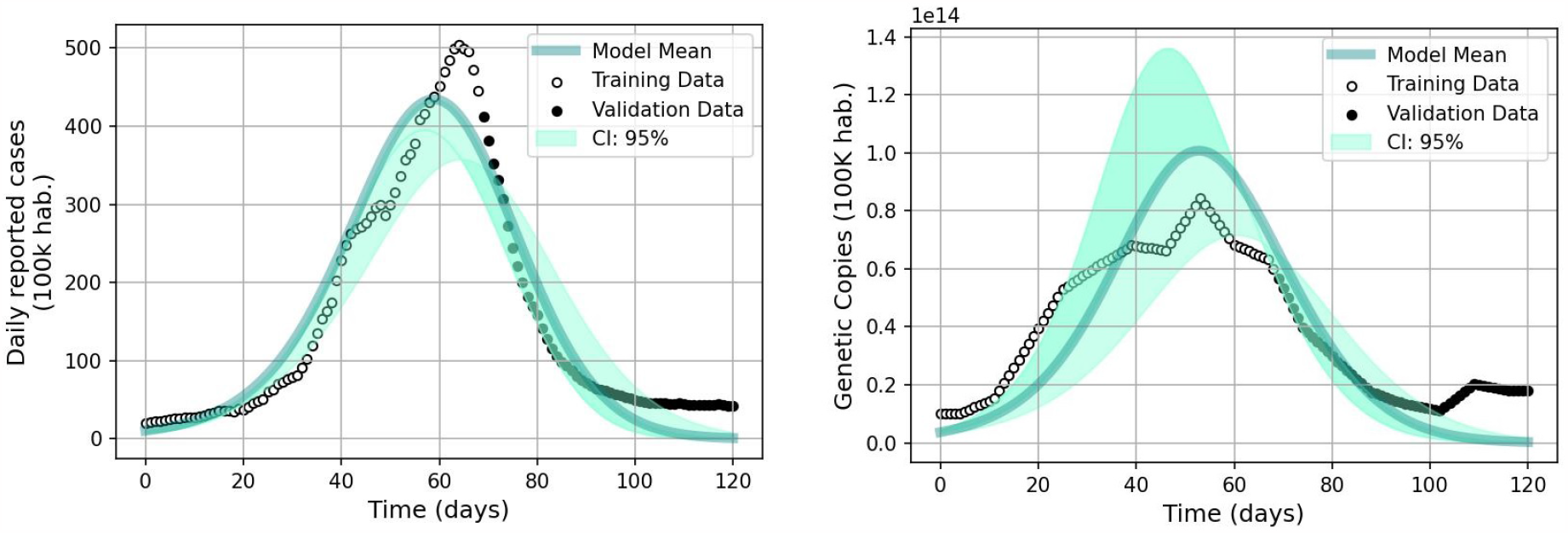
Model validation and spatiotemporal propagation of COVID-19 across Catalonia visualized through daily reported cases and absolute concentrations of genome copies in sewage.

The figures reveal a significant agreement in both qualitative and quantitative terms for all stages of the epidemic wave, particularly in the case of reported cases. Sewage data, which are subject to notable fluctuations, show a lesser degree of agreement.

### 3.3 Detection rate and under-reporting

Our study indicates that the actual number of infections during the period of October 2021 to February 2022 in the analyzed areas of Catalonia was approximately three times higher than the reported cases. However, during November to December 2021, this ratio reached values up to ten (left panel of figure 6). As a result, the detection rate, which is represented by *p*(*t*) in the equations, appears to be a monotonically increasing function over time (central panel in figure 6).

**Figure 6:**
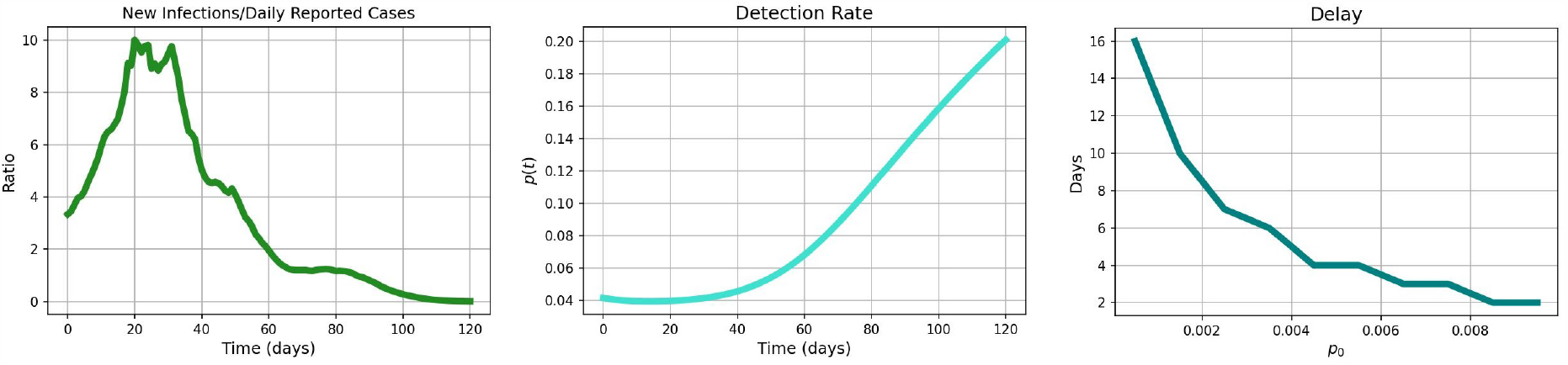
Temporal evolution for the ratio between simulated new infections and daily reported cases data (left panel) and for the transition probability *p*(*t*) (central panel). Delay between generated genome copies in sewage and detected infections versus the parameter *p*_0_.

The model predicted that the daily genome copies would peak approximately five days before the simulated detected infections, which is consistent with some findings in previous literature [6] [8]. This suggests that the observed delays can be attributed to two factors: (i) fluctuations and noise in sewage data, and (ii) the value of the parameter *p*_0_, which is related to the initial value and variability of the detection rate over time. The delay between simulated genome copies and reported cases was observed to be a monotonically decreasing function with *p*_0_ in the equations, with values between 0.001 and 0.01 resulting in a wide range of delays (2 - 16 days), consistent with numerous available datasets (right panel of figure 6).

### 3.4 Maximum quantity of genome copies shed in feces by an individual

Our theoretical framework provides an estimate of the parameter 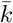 (see Section 2.5) that relates the viral load introduced into the system to that being measured. Using the deterministic equation 2, we estimated *Q*_*max*_, which represents the maximum quantity of genome copies shed in a gram of feces by an individual during the course of infection. This quantity is of interest in the field of Wastewater-Based Epidemiology (WBE) applied to SARS-CoV-2 but has large fluctuations in estimations available in the literature.

Figure 7 shows a colormap indicating the values of *Q*_*max*_ inferred from equation 2 using the mean value of 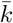 in the posterior distribution yielded by the model calibration procedure. We considered the possible range of values for the dissipation factor *D* (0.86-0.98) and the fraction of people shedding virus in feces *p* (0.29-0.83) as indicated in section 2.5. The results indicate a value of *Q*_*max*_ between 4.15 *×* 10^7^ *gc/g* and 1.33 *×* 10^8^ *gc/g*, which is in agreement with most of the indications from other studies.

**Figure 7:**
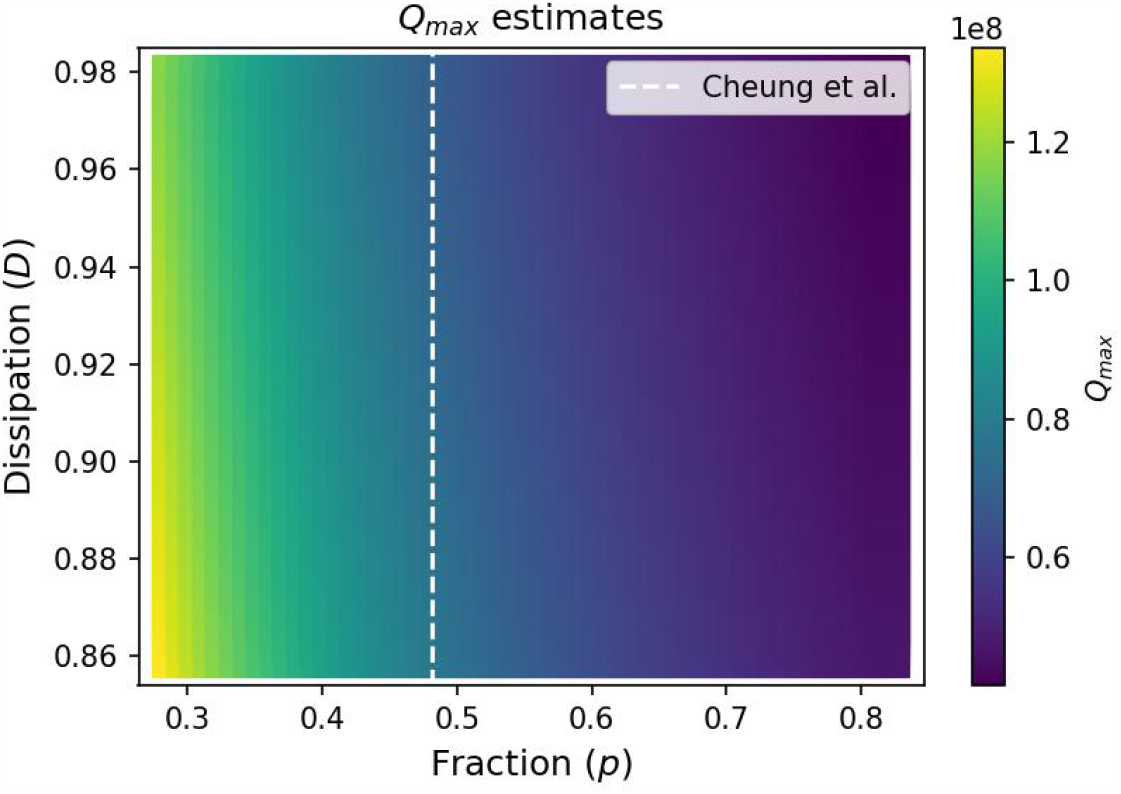
Values of *Q*_*max*_ varying the dissipation factor *D* and the fraction *p* and inferred by the estimated value for 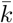.

## 4 Discussion

The aim of this study was to assess the potential of using wastewater-based epidemiology (WBE) to anticipate reported cases and estimate the actual prevalence of SARS-CoV-2 infections. We have used real data from Catalonia. The results showed that wastewater data displayed a high correlation with reported cases, indicating that WBE can capture the current trend of the epidemic. On average, wastewater data anticipated reported cases by about 9-10 days, providing an early warning of an increase in cases.

We have also proposed a simple theoretical framework that integrated wastewater data into a compartmental epidemic model. This framework enabled them to estimate the actual prevalence of infection, which was found to be about 53%, compared to the 19% detected in the same period in Catalonia. This discrepancy suggests that there was a large and time-variable under-reporting in the detection of infections, especially at the onset of the epidemic. We argued that this under-reporting was fundamentally related to people’s perception of the epidemic state and the information available to them, generating a vicious circle.

We have estimated the maximum quantity of genome copies shed in a gram of feces by an individual during the course of the infection, which results to be between 4.15 × 10^7^ *gc/g* and 1.33 × 10^8^ *gc/g*, which showed a good agreement with the literature.

We want also to remark few aspects that can limit our analysis. The main limitation is represented by the data itself: as pointed out by [20], with less than three samples per week the measurements of genome copies in sewage can change according to the day of the data taking. We are looking forward to improve our weekly data-set increasing the number of samples per week. In general, we are aware that a more complex model is needed to model SARS-CoV-2 epidemic involving other aspects like mobility, protection measures, restrictions, age stratification etc.. and, in particular, to express such intricate concept like the people perception and awareness about the epidemic. Indeed, other aspects of human behaviour can be taken in consideration, as imitation processes or adoption of different strategies, given that human behaviour and epidemic spreading undergo to a complex interaction that goes in both directions. We are also aware that mechanistic models that try to in-globe wastewater data cannot be extremely accurate, due to the intrinsic volatility and the multitude of factors that enter in the entire process of the virus shedding in the sewage system. For instance, Thomas et al. [34] highlighted how considering dynamical populations, for which the number of persons served by a specific sewage plant can change in time, is way more accurate than fixed ones. Morvan et al. [8] showed how machine learning models result naturally more accurate in capturing the wastewater phenomenology.

Nevertheless, we think that our work can hopefully highlight the importance of monitoring trends in wastewater data, being a crucial tool to estimate actual infections when voluntary testing policies results too biased. We hope that this work can be seen as a threefold contribution in the field of wastewater-based epidemiology, in the study of biases in data and as a retrospective study for the Covid-19 Omicron epidemic wave in Catalonia during January 2022.

## Data Availability

All data analyzed are available online

https://sarsaigua.icra.cat/

https://analisi.transparenciacatalunya.cat/browse?q=covid&sortBy=relevance

## Acknowledgments

This work was partially funded by the Catalan Agency for Water (ACA), the Catalan Public Health Agency (ASPCAT) from the Department of Health, and the Health Innovation Program from the General Research Directorate (DGRIS) of the Generalitat de Catalunya. The authors would like to thank the EU and The Spanish State Research Agency for funding project PCI2021-121928, in the frame of the collaborative international consortium SARA financed under the ERA-NET AquaticPollutants Joint Transnational Call (GA n^a^869178). We kindly acknowledge Promega for the assignment of a Maxwell AS4500 nucleic acids extraction System.

This project has received funding from the European Union’s Horizon 2020 research and innovation program under the Marie Sklodowska-Curie grant agreement No. 945413 and from the Universitat Rovira i Virgili (URV). Disclaimer: This work reflects only the author’s view and the Agency is not responsible for any use that may be made of the information it contains.

AA acknowledges the Joint Appointment Program at Pacific Northwest National Laboratory (PNNL). PNNL is a multi-program national laboratory operated for the U.S. Department of Energy (DOE) by Battelle Memorial Institute under Contract No. DE-AC05-76RL01830, the European Union’s Horizon Europe Programme under the CREXDATA project, grant agreement no. 101092749, support by Ministerio de Economía y Competitividad (Grants No. PID2021-128005NB-C21), Generalitat de Catalunya (Grant No. 2017SGR-896) and Universitat Rovira i Virgili (Grant No. 2019PFR-URV-B2-41). A.A. also acknowledges ICREA Academia 2023, and the James S. McDonnell Foundation (Grant No. 220020325).

## Authors contribution

M.M. and A.A. designed the study. R.P., S.G., and A.B. provided the data. M.M. analyzed the data and performed the analysis. M.M. and A.A. analyzed the results and wrote the paper. All authors reviewed and approved the complete manuscript.

## Figure captions

- **Figure 1**: Histogram showing the distribution of the delays between wastewater data and reported cases curve across the 16 WWTPs considered. With *delay* we refer to the amount of days by which daily reported cases needed to be shifted back in time in order to obtain maximum Pearson correlation. This procedure results in a average correlation of 0.88*±*0.08 (0.71-0.96) and an average delay of 8.7 *±* 5.4 days (0-20);
- **Figure 2**: Graphic showing the trends for absolute genome copies concentrations of SARS-CoV-2 averaged across the 16 WWTPs (dots, right y-axis) and for 7-days averaged reported cases in Catalonia (solid line, left y-axis). Both measures were calibrated to 100.000 inhabitants. The striped area indicates the temporal distance between the peaks of the two curves;
- **Figure 3**: The figure shows the results of the Approximate Bayesian Computation (ABC) procedure for the estimation of the parameters *β, p*_0_ and 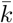, respectively indicated as *b, p* and *k*. The plots on the left side are the posterior distributions whereas in the right side are showed all the values sampled during the process for all three parameters;
- **Figure 4**: The figure displays the temporal evolution of the simulated epidemic according to the model. The curves indicates the proportion of susceptible (S), infected not detected (*I*_*N*_), infected detected (*I*_*D*_) and recovered (*R*) compartments;
- **Figure 5**: The figure shows the comparison between model predictions and data about daily reported cases (left side) and absolute genetic concentrations of SARS-CoV-2 in sewage. Solid lines show model predictions for the daily reported cases (left) and the daily number of genome copies in sewage (right) for 100 000 inhabitants, whereas dots correspond to real data. The model has been trained for the first 100 days data (white dots) and validate in the remaining ones (black dots). The shadowed areas represent the 95% prediction interval. The *R*^2^ statistics is 0.94 for cases and 0.64 for wastewater data;
- **Figure 6**: Left panel: temporal evolution of the ratio between new infections simulated by the model at each time step and daily reported cases data. Central panel: temporal evolution of the detection rate *p*(*t*) according to the model estimates. Right panel: days of delay between generated quantity of genome copies concentrations and detected infections varying the value of *p*_0_. The former is deduced again looking to the maximum Pearson correlation between the two simulated data-sets of genome copies and detected infections;
- **Figure 7**: Colormap showing estimations for the maximum quantity of genome copies shed in feces by an individual *Q*_*max*_ inferred from equation 2, varying the dissipation factor (*D*) and the fraction of people shedding virus in feces (*p*), according to the indications in the literature. We used the mean value of 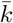 in the posterior distribution yielded by the model calibration procedure. The white dashed line indicates the value for *p* suggested by Cheung et al. [24] in their review.

